# Demographics of COVID19 vaccine hesitancy during the second wave of COVID-19 pandemic: A cross-sectional web-based survey in Saudi Arabia

**DOI:** 10.1101/2021.04.20.21255781

**Authors:** Mohammed Al-Mohaithef, Bijaya Kumar Padhi, Soukaina Abdulmajed Ennaceur

## Abstract

**Background:** The Coronavirus disease 2019 (COVID-19) pandemic is considered a major global public health threat affecting across the life course and socioeconomic aspects of life. Globally acceptance to an effective vaccine is the most anticipated resolution. This study aims to evaluate intent to be vaccinated among public in Saudi Arabia during the second wave of COVID-19 pandemic.

**Methods:** A cross-sectional web-based study was designed in Saudi Arabia. Study participants (N=658) were recruited through snowball sampling. SurveyMonkey platform was used to record the response. Cross-tabulation were performed by participants’ intention to vaccinate against COVID-19 virus with sociodemographic characteristics and respondents’ risk perception towards COVID-19, trust in the healthcare system, and their history of vaccine hesitancy behavior. Multivariable logistic regression analysis was performed to compute the predictors of vaccination intention among the study participants.

**Results:** 658 participants were completed the survey (females = 47.4%). Of the 658 participants 351 (53.3%) have shown intent to be vaccinated. 519 (78.8%) of the participants were reported to be at high risk of COVID-19, and 307 (46.6%) were reported to trust the healthcare system in the country. The multivariable analysis shows respondents with a high-risk perception (OR: 2.27, 95% CI: 1.49-3.48); higher trust in the healthcare system (OR: 3.24, 95% CI: 2.32-4.61) was found to be the significant factor affecting the decision in acceptance of the COVID-19 vaccine in Saudi Arabia.

**Conclusion:** Participants reported high knowledge towards COVID-19 virus, and vaccine developments. About half (46.6%) of the study participant reported refusal/hesitancy towards the vaccine during the second wave of the pandemic in Saudi Arabia. The study highlighted that higher risk perception and higher trust in the healthcare system were found to be the main reasons for participant’s intentions behind the vaccination.

## Introduction

During the preceding year 2019-2020, all the countries worldwide were affected by a novel severe acute respiratory syndrome coronavirus-2 named the COVID-19, which became an international concern. By February 2021, the World Health Organization (WHO) has declared more than 108 M confirmed cases of COVID-19 and 2.3 M deaths all over 219 countries and territories(WHO, 2021). Numerous studies have shown concern for the second wave of COVID-19(Nicholson & Krings, 2021; Xu & Li, 2020).

As vaccination is considered the most effective health intervention to prevent and control the COVID-19, scientists worldwide competed to develop a safe and productive vaccine in record time. By November 2020, the German BioNTech, in cooperation with the US-Pfizer, announced that their vaccine has a more than 90% success rate. The clinical trial analysis showed that this vaccine caused no severe side effects, resulting in rapid use across the world(WHO, 2021). In the middle of February 2021, WHO declared at least seven different COVID-19 vaccines have been rolling out in other countries. Moreover, 200 additional vaccines are in development, of which more than 60 are in clinical outcome(WHO, 2021).

The Kingdom of Saudi Arabia (KSA) was one of the very first countries in the world to grant emergency use authorization to the BNT162b2 vaccine(Barry & BaHammam, 2021) and received the first batch of the COVID-19 vaccine by December 2020(Ministry of Health Saudi Arabia, 2020a). One month later, over one million people were registered on the local application “Sehhaty” to receive the vaccine(Ministry of Health Saudi Arabia, 2020c). By 15 April, the Ministry of Health announced around three million people have taken the vaccine shot without any health complications. The COVID-19 vaccine is provided free of charge to all Saudi Arabia (Saudi and non-Saudi) inhabitants, and priority is given to those with chronic diseases, health practitioners, and individuals over 65 years(Ministry of Health Saudi Arabia, 2020b).

The acceptance of a vaccine is a critical stage in the success of the immunization programs to accomplish high vaccination rates within the general population. The acceptance of the vaccine depends largely on many factors such as the way people perceive the threat, the thrust on the healthcare system, the vaccine safety and efficacy, and the vaccine demand within the population(Wilson et al., 2011; Yang et al., 2020). Many studies on the acceptance and the confidence in the vaccines, mainly for the newly emerging infectious diseases, showed inadequate acceptance rates. For example, during the 2009 H1N1 pandemic, it has been demonstrated that in France, the acceptance rate of the vaccine was 17%; in America, 49.6%; in Australia 43.9%; in the UK 56.1%; and in Greece, the rejection rate of the vaccine was 63.1% (Maurer et al., 2010; Schwarzinger et al., 2010; Seale et al., 2010; Sypsa et al., 2009). A recent study conducted in Saudi Arabia to assess the prevalence of vaccine hesitancy or delay among the Saudi population showed that 36% of children are not vaccinated fully at their age(Alsubaie et al., 2019). The study reported that children’s parents believed vaccination was ineffective or unnecessary and thought that receiving a vaccine for diseases that are no longer common is insignificant (Alsubaie et al., 2019).

A global survey of 19 countries reported that about 71.5% of participants would be very or somewhat likely to take a COVID-19 vaccine(Lazarus et al., 2020a). A systematic review highlighted that several countries reported the lowest COVID-19 vaccine acceptancy, which includes Kuwait (23.6%), Jordan (28.4%), Italy (53.7), Russia (54.9%), Poland (56.3%), US (56.9%), and France (58.9%). A study conducted among the Middle Eastern population showed that 36.8% and 26.4% of the participants answered “No” and “Not sure” when asked if they would take the vaccine once it becomes available(Al-Qerem & Jarab, 2021). In Saudi Arabia, a study was conducted before the new COVID-19 vaccine approval and authorization of the use by our research group and aimed to understand the public willingness of a future COVID-19 vaccine. The findings showed that a high proportion (64.7%) of the study participants demonstrated intention and desire to receive the vaccine(Al-Mohaithef & Padhi, 2020) [16]. About half (46.7%) of the young adult population reported taking the COVID-19 vaccine if made mandatory in Saudi Arabia (Almaghaslah et al., 2021). Another study showed 50.52% of healthcare workers were willing to have the COVID-19 vaccine. However, 50.29% intended to delay until the vaccine’s safety is confirmed(Qattan et al., 2021). However, the concrete rate of hypothetical vaccine acceptance might be much inferior to the acceptance rate after the development and authorization of vaccine use through a national immunization program. The present study was conducted to assess the COVID-19 vaccine acceptance among the Saudi population and determine the demographic characteristics that might impact the vaccine uptake during the second wave of the pandemic.

## Methods

### Study Design and Sample

A cross-sectional survey was conducted on the acceptability of a COVID-19 vaccine among the public between January 2021 to March 2021 in Saudi Arabia. The OpenEpi 3.01 (updated on April 2013) was used for the calculation of sample size considering the anticipated proportion of COVID-19 vaccine acceptance 40% obtained from the recent studies(Al-Mohaithef & Padhi, 2020; Almaghaslah et al., 2021; Qattan et al., 2021); with absolute precision of 5% and 99% confidence interval. The sample size was estimated to be 637. Study participants were recruited through snowball sampling. Invitations to participate in the study were distributed to the respondents via Email, social media: Twitter, Facebook, and the WhatsApp communication platform of the primary contacts of the study team and requested to transmit further for its maximum reach. Considerations were made to recruit the participants across major geographic regions of the KSA.

### Measures

The development and validation of survey questionnaires were reported elsewhere(Al-Mohaithef & Padhi, 2020). Briefly, a bilingual self-reported format questionnaire was developed using SurveyMonkey. Informed consent was obtained from all participants (18 years or older and currently living in the KSA). Detailed responses were recorded further only those who provided informed consent to complete the survey.

The first section of the questionnaire includes information on sociodemographic characteristics of study participants such as age, gender, marital status, education level, place of residency, occupation sector, socio-economic status. The second section collects information on the history of exposure to COVID-19 (travel with a confirmed COVID-19 patient, staying with a confirmed COVID-19 patient); risk perception; concerns to be affected by COVID-19; trust in the healthcare system. The third section of the questionnaire records information on knowledge about the development of the COVID19 vaccine and intention to be vaccinated against the COVID-19 virus.

### Statistical Analyses

The key outcome measures of the study were to know the respondent’s intention towards acceptance of the COVID-19 vaccination. The responses were recorded in a three-point Likert scale (Yes, No, and Not sure). Descriptive statistical analysis was performed by doing cross-tabulation of demographic characteristics with the primary response variable. The variable ‘age’ was converted into three categories: 18–25 (the reference category), 26-35, and above 35 years. Gender was reported in two categories: male and female. Marital status was presented in two categories: married and single (including widowed and divorced). Education status was captured into four categories: high school, diploma, undergraduate, and postgraduate, and above. Participants’ socio-economic status (SES) was reported into three categories: low, medium, and high. The family size was reported into two categories: ≤5 and ≥6. The place of residence was captured numerically, covering all 13 administrative regions in the KSA (Riyadh, Mekkah, Almadina Almonawra, Qaseem, Eastern Region, Aseer, Tabouk, Hail, Northern Borders, Jazan, Najran, Albaha, and Aljouf), and further grouped into five categories: East, West, North, South, and Central.

We performed both simple and multivariable logistic regression analyses to compute the odds ratio (OR) and a 95% confidence interval (CI). Chi-squared tests were performed for bivariate analysis (cross-tabulation) between the outcome variable (intend to vaccinate) and all explanatory variables. Inference on significant association was considered with a two-tailed p≤0.05. STATA 15.0 software (StataCorp LP, Texas, USA) was used for all statistical analysis.

### Ethical Considerations

Ethical approval was granted for the study by the institutional Research Ethics Committee (SEUREC-CHS20110) Saudi Electronic University, Riyadh, Kingdom of Saudi Arabia, and consent was taken before participation in the study. Anonymized data was used for analysis, interpretation, and reporting.

## Results

Table 1 shows the demographic characteristics of the study participants. Of the 658 participants 286 (43.4 %) were aged 26 to 35 years old; 312 (47.4%) were female; and 326 (49.5%) were married respondents. The majority of respondents belonged to the central provinces, 288 (43.7%), followed by western 200 (30.3%), and 170 (25.8%) eastern provinces in Saudi Arabia. The most common education level was undergraduate and below (80.2%), and the majority (n=413) were employed in governmental or private sectors or self-employed. Only 15.19% worked in the health care system (hospitals, clinics, and laboratory testing and diagnosis). High responses were from respondents living in large-sized families (six children and above) (66.4%) compared to low-sized families (five children and below) (37.4%). The study has high participation of respondents with medium social-economic status (81.4%).

Table 2 presents contact history with COVID-19 patients, risk perception, and vaccination history of the study respondents. In terms of contact history with COVID-19 patients, a low proportion of respondents stated that they traveled (6.84%) or stayed (19.00%) in the same environment with a confirmed COVID-19 patient. The respondents had a high-risk perception of the COVID-19 (78.88%) and agreed that they are concerned and afraid of getting the coronavirus (79.78%). 46.6% of the respondents reported high trust in the healthcare system in Saudi Arabia to manage the current situation related to COVID-19, but 53.44 % showed low trust in the healthcare system. Regarding the history of vaccine hesitancy, 81.0% disagreed with receiving one or more types of vaccine if there were doubts about its efficacy and safety. While 18.99 % refused to accept any vaccination and considered it without any benefit to their health and their children’s health. 14.28% declared that they postponed a vaccine even if their physician recommended it.

Table 3 depicts the bivariate analysis for the association between demographics characteristics and intention to COVID-19 vaccination. We conducted a cross-tabulation analysis to examine the distribution of intentions to uptake the COVID-19 vaccine according to the sociodemographic characteristics of the respondents. In general, 53.35% (n= 351) of the respondents declared positive intention to update COVID-19 vaccine, while 46.65 % (n= 307) responded “No or Not sure” to receive the vaccine. The cross-tabulation analysis using the Chi-square test for binary or categorical variables showed that a statistically significant number of married respondents (53.85%) declared a strong intention to receive the COVID-19 vaccine when compared to the unmarried respondents (46.55 %) (*p*= 0.02). Moreover, by education levels, a significantly higher proportion of respondents who expressed an absolute intent to vaccinate included respondents with undergraduate education (45.30%) or postgraduate respondents (24.22%) (*p*= 0.01). There was also significant intention to update the COVID-19 vaccine among highly sited families with six children or more (59.26%) when compared to families with less than five children (40.74%) (*p*=0.06). Respondents aged between 26 and 35 years old (41.03%), male respondents (54.99%), and respondents with medium socio-economic status (82.62 %) also declared strong and definite intention to vaccination when compared to the other categories from the same class; however, these differences did not show any statistical significance (*p*>0.05).

Table 4 shows the bivariate analysis for the association between intention to COVID-19 vaccination with vaccine knowledge, trust, and confidence in the coronavirus vaccination among the study respondents. Regarding the knowledge about the development of the COVID-19 vaccine, 76.07% of respondents with high levels of awareness confirmed high trust and confidence in the vaccination. In comparison, 54.07% showed no preference for the COVID-19 vaccine despite their knowledge about the importance and development of the vaccine (*p*<0.001). When the respondents were asked if they trust the healthcare system in Saudi Arabia, 66.38% responded “Yes,” which could impact positively the trust and confidence in the COVID-19 vaccine (*p*<0.001) when compared with respondents who showed no trust in the healthcare system (38.4%).

Moreover, a significant proportion thought that domestic vaccines are worse than imported vaccines (46.9%), affecting their trust in the COVID-19 vaccination, while only 19.37% trusted the domestic vaccines. There was also a statistically significant difference between respondents with a high-risk perception of the COVID-19 (85.02%) but declared no trust and confidence to receive the vaccine when compared to 73.50 % of respondents with a high perception of the risk and who showed significant faith and enthusiasm to the COVID-19 vaccine (*p*<0.05). The history of vaccination hesitancy, the history of contact with confirmed COVID-19 patients, and the exposure to the COVID-19 were without any impact on the trust and confidence in the COVID-19 vaccine (*p*>0.05).

Table 5 shows a multivariable logistic regression analysis of the factors influencing the COVID-19 vaccine uptake among study participants. As the respondents were divided into two groups according to their intention to vaccinate (53.35 % “Yes” vs. 46.65 % “No or Not sure”), a multivariable logistic regression analysis was then conducted between these two groups to identify factors that may impact the acceptance or refusal of the vaccine uptake. Basic sociodemographic characteristics, history of vaccine hesitance, risk perception, and trust in domestic vaccinations were studied, with the refusal group of the vaccine considered as the reference group (Table 5). Respondents with a high perception of vaccination benefit in reducing the spread of the disease and control the risk of the COVID-19 are showing high intention to be vaccinated (OR: 2.27, 95% CI: 1.49-3.48). The trust in the healthcare system and confidence in its role to manage the situation in Saudi Arabia and to prevent and control the COVID-19 (OR: 3.24, 95% CI: 2.32-4.61) was found to be the significant factor affecting the decision making of the respondents regarding the acceptance of the COVID-19 vaccine.

## Discussions

In this study, we aimed to determine the acceptability and hesitancy towards vaccination during the second wave of COVID-19 pandemic in Saudi Arabia, and the factors associated with these issues. We found that nearly 53.3% of adults in Saudi Arabia would be willing to receive a COVID-19 vaccine vs 64.7% during the first phase of the pandemic(Al-Mohaithef & Padhi, 2020). Sociodemographic factors, such as being married, and higher educational level, were found to be associated with vaccine acceptance. Higher risk perception and higher trust in the healthcare system were found to be the significant predictors in explaining participants’ intentions behind the vaccination. Our findings represent one of the first estimates of the acceptance of a COVID-19 vaccine in Saudi Arabia during the second wave of COVID-19 pandemic.

Even though this study was conducted between January-March 2021 when the second wave of COVID-19 approaching and the country is ready with vaccine to immunize the community, only 53.3% of participants intend to be vaccinated. This percentage is comparable lower than our previous finding when there was no vaccine available in the country(Al-Mohaithef & Padhi, 2020). But still the community in Saudi Arabia have a higher rate of acceptance towards the vaccine in comparison to the countries like Kuwait (23.6%), and Jordan (28.4%) (Lazarus et al., 2020b). Even the general public had a higher acceptability to the vaccine than healthcare workers in Saudi Arabia(Qattan et al., 2021). The healthcare workers reported 50.52% willingness to have the COVID-19 vaccine in Saudi Arabia(Qattan et al., 2021). The young adults have a lower acceptability (48.0%) of the vaccine in Saudi Arabia as reported in a recent study (Almaghaslah et al., 2021).

Our study also highlighted that public trust on healthcare system and risk perception towards the virus were the significant predictive behavior in accepting the vaccine as stated in previous studies(Almaghaslah et al., 2021; Alqudeimat et al., 2021; Barello et al., 2020; Machida et al., 2021; Qattan et al., 2021, 2021).

The strength of our study includes an appropriate sample size and recruitment of study participants across the country. The limitation of this study includes sampling strategy, we used a snowball sampling which may have been overestimated due to selection bias. Secondly, we did not account some psychological factors which may influence the acceptance of vaccination. Despite these limitations, to the best of our knowledge, this is the first study to report the current COVID-19 vaccine acceptance in the country while there is a vaccine is available and the second wave of pandemic devastating the community.

## Conclusion

This study depicts that the study participants have good knowledge of COVID-19 and vaccine developments. However, the participants’ vaccination intentions were only 53.3% during the second wave of the pandemic. The study highlighted that higher risk perception and higher trust in the healthcare system were found to be the main reasons for participants’ intentions behind the vaccination. Policymakers should priorities these concerns by creating awareness among the community.

## Data Availability

The data is available with the corresponding authors and provided up on request

http://Seu.Riyadh

## Notes

### Competing Interest Statement

The authors have declared no competing interest.

### Funding Statement

Nil

