## Supplementary material for "Demographics of COVID19 vaccine hesitancy during the second wave of COVID-19 pandemic: A cross-sectional web-based survey in Saudi Arabia": Table 1-5

**Table Captions**

**Table 1:** Socio-demographic characteristics of the study respondents (N=658)

**Table 2:** Contact history with COVID-19 patients, risk perception, and vaccination history of the study respondents (N=658)

**Table 3:** Bivariate analysis for association between demographics characteristics and intention to COVID-19 vaccination (N=658)

**Table 4**: Bivariate analysis for association between intention to COVID-19 vaccination with vaccine knowledge, trust, and confidence among the study respondents (n-658)

**Table 5:** Multivariable Regression Analysis of the influencing factors on the COVID-19 vaccine uptake among study participants

**Table 1:** Socio-demographic characteristics of the study respondents (N=658)

| Variables | Categories | n (%) |
| --- | --- | --- |
| Age | 18-25 | 189 (28.72%) |
|  | 26-35 | 286 (43.47%) |
|  | >35 | 183 (27.81%) |
| Gender | Male | 346 (52.58%) |
|  | Female | 312 (47.42%) |
| Marital status | Single | 332 (50.46%) |
|  | Married | 326 (49.54%) |
| Highest education | High School | 120 (18.24%) |
|  | Diploma | 94 (14.29%) |
|  | Undergraduate | 314 (47.72%) |
|  | Postgraduate | 130 (19.76%) |
| Employment status | Governmental sector | 244 (37.08%) |
|  | Private sector | 142 (21.58) |
|  | Self-employed | 27 (4.10) |
|  | Unemployed | 245 (37.23) |
| Socio-economic status | Low | 77 (11.70%) |
|  | Medium | 536 (81.46%) |
|  | High | 45 (6.84%) |
| Family size | ≥6 members | 412 (62.61%) |
|  | ≤5 members | 246 (37.39%) |
| Place of residence | Eastern region | 170 (25.83%) |
|  | Central region | 288 (43.76%) |
|  | Western region | 200 (30.39%) |
| Working in the health care system | Yes | 100 (15.19%) |
|  | No | 588 (89.36%) |

**Table 2:** Contact history with COVID-19 patients, risk perception, and vaccination history of the study respondents (N=658)

| Variables | Categories | n (%) |
| --- | --- | --- |
| History of travel with a confirmed COVID-19 patient | Yes | 45 (6.84%) |
|  | No | 613 (93.16%) |
| History of staying with a confirmed COVID-19 patient | Yes | 125 (19.00%) |
|  | No | 533 (81.00%) |
| Risk perception | Yes | 519 (78.88%) |
|  | No | 139 (21.12%) |
| Concerns to be affected by  COVID-19 | Yes | 525 (79.78%) |
|  | No | 133 (20.21%) |
| Trust in the healthcare system | Yes | 307 (46.66%) |
|  | No | 351 (53.34%) |
| Uptake other types of vaccine despite doubts about their efficacy | Yes | 120 (18.23%) |
|  | No | 538 (81.76%) |
| Refused other types of vaccines | Yes | 125 (18.99%) |
|  | No | 533 (81.00%) |
| Postpone a vaccine recommended by a physician | Yes | 564 (85.71%) |
|  | No | 94 (14.28%) |

**Table 3:** Bivariate analysis for association between demographics characteristics and intention to COVID-19 vaccination (N=658)

| Variables |  | No/Not sure  (n=307; 46.65%) | Yes  (n = 351; 53.35%) | Total  (n=658) | *p*-value |
| --- | --- | --- | --- | --- | --- |
| Age | 18-25 | 91 (29.64%) | 98 (27.92%) | 189 (28.72%) | 0.13 |
|  | 26-35 | 142 (46.25%) | 144 (41.03%) | 286 (43.47%) |  |
|  | above 35 | 74 (24.10%) | 109 (31.05%) | 183 (27.81%) |  |
| Gender | Male | 153 (49.84%) | 193 (54.99%) | 346 (52.58%) | 0.19 |
|  | Female | 154 (50.16%) | 158 (45.01%) | 312 (47.42%) |  |
| Marital status | Single | 170 (55.37%) | 162 (46.15%) | 332 (50.46%) | 0.02 |
|  | Married | 137 (44.63%) | 189 (53.85%) | 326 (49.54%) |  |
| Highest education | High School | 64 (20.85%) | 56 (15.95%) | 120 (18.24%) | 0.01 |
|  | Diploma | 43 (14.01%) | 51 (14.53%) | 94 (14.29%) |  |
|  | Undergraduate | 155 (50.49%) | 159 (45.30%) | 314 (47.72%) |  |
|  | Postgraduate | 45 (14.66%) | 85 (24.22%) | 130 (19.76%) |  |
| Socio-economic status | Low | 41 (13.36%) | 36 (10.26%) | 77 (11.70%) | 0.46 |
|  | Medium | 246 (80.13%) | 290 (82.62%) | 536 (81.46%) |  |
|  | High | 20 (6.51%) | 25 (7.12%) | 45 (6.84%) |  |
| Family size | Six and above | 204 (66.45%) | 208 (59.26%) | 412 (62.61%) | 0.06 |
|  | Five and below | 103 (33.55%) | 143 (40.74%) | 246 (37.39%) |  |

**Table 4**: **Bivariate analysis for association between intention to COVID-19 vaccination with vaccine knowledge, trust, and confidence among the study respondents (n-658)**

| Variables | No/Not sure | Yes | Total (658) | *p*-value |
| --- | --- | --- | --- | --- |
| **History of travel with a confirmed COVID-19 patient** |  |  |  |  |
| No/Not Sure | 284 (92.51%) | 329 (93.73%) | 613 (93.16%) | 0.53 |
| Yes | 23 (7.49%) | 22 (6.27%) | 45 (6.84%) |  |
| **History of staying with a confirmed COVID-19 patient** |  |  |  |  |
| No/Not Sure | 255 (83.06%) | 278 (79.20%) | 533 (81.00%) | 0.21 |
| Yes | 52 (16.94%) | 73 (20.80%) | 125 (19.00%) |  |
| **Exposed to COVID-19 cases** |  |  |  |  |
| No | 243 (79.15%) | 276 (78.63%) | 519 (78.88%) | 0.87 |
| Yes | 64 (20.85%) | 75 (21.37%) | 139 (21.12%) |  |
| **Knowledge about COVID19** |  |  |  |  |
| No/Not Sure | 65 (21.17%) | 19 (5.41%) | 84 (12.77%) | 0.00 |
| Yes | 242 (78.83%) | 332 (94.59%) | 574 (87.23%) |  |
| **Knowledge about the development of the COVID19 vaccine** |  | |  |  |
| No/Not Sure | 141 (45.93%) | 84 (23.93%) | 225 (34.19%) | 0.00 |
| Yes | 166 (54.07%) | 267 (76.07%) | 433 (65.81%) |  |
| **History of vaccine hesitancy** |  | |  |  |
| Yes | 67 (21.82%) | 86 (24.50%) | 153 (23.25%) | 0.42 |
| No | 240 (78.18%) | 265 (75.50%) | 505 (76.75%) |  |
| **Risk perception** |  |  |  |  |
| Yes | 261 (85.02%) | 258 (73.50%) | 519 (78.88%) | 0.00 |
| No | 46 (14.98%) | 93 (26.50%) | 139 (21.12%) |  |
| **Trust in the healthcare system** |  |  |  |  |
| No | 189 (61.56%) | 118 (33.62%) | 307 (46.66%) | 0.00 |
| Yes | 118 (38.44%) | 233 (66.38%) | 351 (53.34%) |  |
| **Vaccine brand (domestic)** |  |  |  |  |
| Better | 45 (14.66%) | 68 (19.37%) | 113 (17.17%) | 0.00 |
| Similar | 55 (17.92%) | 111 (31.62%) | 166 (25.23%) |  |
| Worse | 144 (46.91%) | 137 (39.03%) | 281 (42.71%) |  |
| Not Sure | 63 (20.52%) | 35 (9.97%) | 98 (14.89%) |  |

**Table 5:** Multivariable Regression Analysis of the influencing factors on the COVID-19 vaccine uptake among study participants

| Variable | OR [95% CI] | *p*-value |
| --- | --- | --- |
| History of vaccine hesitancy | 0.74 [0.50 - 1.09] | 0.13 |
| Risk perception | 2.27 [1.49 - 3.48] | 0.00 |
| Trust in the healthcare system | 3.24 [2.32 - 4.51] | 0.00 |
| Trust in domestic vaccines | 1.26 [0.90 - 1.76] | 0.17 |
| Age | 0.93 [0.70 - 1.25] | 0.64 |
| Gender | 0.72 [0.51 - 1.02] | 0.07 |
| Marital status | 1.21 [0.80 - 1.83] | 0.37 |
| Highest education | 1.16 [0.97 - 1.40] | 0.10 |
| Socio-economic status | 1.16 [0.78 - 1.72] | 0.46 |
